# Effect of Lumbar Medial Branch Nerve Radiofrequency Ablation on Lumbar Spondylolisthesis

**DOI:** 10.1101/2020.11.10.20227900

**Authors:** Trusharth Patel, Christopher Watterson, Anne Marie McKenzie-Brown, Boris Spektor, Katherine Egan, David Boorman

## Abstract

**Importance:** Radiofrequency ablation (RFA) is a denervation therapy commonly performed for pain of facet etiology. Degenerative spondylolisthesis may be a co-existing condition; yet the effect of RFA on advancing listhesis is unknown.

**Objective:** To test the hypothesis that RFA of painful facets in the setting of spondylolisthesis may contribute to advancement of further degenerative spondylolisthesis.

**Design:** Retrospective and prospective, observational study conducted at a single academic center among 15 participants with pre-existing degenerative Grade I or Grade II spondylolisthesis undergoing lumbar RFA encompassing spondylolisthesis level and followed with post-RFA imaging at 12 months and beyond to measure percent change in spondylolisthesis.

**Main Outcomes and Measures:** The primary outcome was the percent advancement of spondylolisthesis per year measured on post-RFA lateral lumbar spine imaging compared to non-intervention baseline advancement of 2.6% per limited observational studies.

**Results:** Among the 15 participants enrolled, 14 completed the study (median age 66; 64.3% women; median BMI 33.5; mean follow-up time 23.9 months). The mean advancement of spondylolisthesis per year after RFA was 1.30% (95% CI −0.14 to 2.78%), with 9/14 below 1.25%.

**Conclusion and Relevance:** Among patients with lumbar pain originating from facets in the setting of degenerative spondylolisthesis who underwent lumbar RFA, the observed advancement of spondylolisthesis is clinically similar to the baseline of 2.6% per year change. The study findings did not find a destabilizing effect of lumbar RFA in advancing spondylolisthesis in this patient population.

## Introduction

Spondylolisthesis is a spine condition which refers to a malalignment of the vertebrae of the spine between two adjacent levels. There are numerous causes of spondylolisthesis; its presence can ultimately lead to pain, spinal stenosis, neuroforaminal stenosis and instability of the spine by loss of a stabilizing mechanism of the articular processes of the vertebrae. Lower back pain can be a clinical presenting sign; however, the etiology and pathomechanics remain unclear.^1^ Degenerative spondylolisthesis is the most common type of spondylolisthesis; the prevalence varies based on the population studied. In the Chinese male and female population, the prevalence is 19% and 25%, respectively.^2^ In the Australian male and female population over the age of 50, the prevalence is 7.5% and 16.7-28.7%, respectively.^3^ In the American female population, epidemiological studies show a prevalence in older white females of 19-29%, and older African American females of 60%.^4.5^ Several risk factors to developing spondylolisthesis have been described: body mass index (BMI), age, and angle of lordosis in women were significantly associated with degenerative spondylolisthesis; increased age was associated in men.^6^ Mean difference in BMI through this study period only varied marginally and the study population was homogenous making generalization to other populations difficult. A systematic review did find overall low to very low quality evidence findings on risk factors associated with lumbar degenerative spondylolisthesis. Increased age, female gender, and increased facet joint angle as potential risk factors show low quality evidence; back pain, prolonged occupational sitting, lumbosacral angle, lumbar lordosis, and parity show even lower quality evidence to support an association.^7^ Position of imaging, recumbent vs. sitting or standing has been proposed as a theoretical association in observing spondylolisthesis; however, neither a large study on an elderly Chinese population nor a 45-year prospective study found this association.^8,9^

The rate of progression of degenerative lumbar spondylolisthesis remains unclear, as are independent risk factors for progression. Progression of slip in children with pars defects seems to be more prominent in early decades and slows significantly in adulthood at the fourth decade of follow-up evaluation.^9^ Degenerative spondylolisthesis is reported to be uncommon prior to age 40 for men and 50 for women.^10^ The prevalence of progression of spondylolisthesis for 300 elderly men at 4.6-year follow up has been reported to be 12%, with advancement of slippage defined as >5%. In the same study, 12% of de novo cases were reported in the same study period.^11^ These results are comparable to what was observed in a 4-year follow-up study of a Chinese sample population of men with spondylolisthesis progressing in 13% of men; de novo cases appeared in 12.4%. In the same study, spondylolisthesis progressed in 16.5%; de novo cases appeared in 12.7% of women.^8^ Neither study quantified the rate of progression of actual slippage over time. One study examined the rate of progression of slippage of vertebrae in 311 patients younger than 30 years old with spondylolisthesis resulting from spondylolysis. Peak slippage in this younger population occurred between the ages of 20-25 years and average slippage per year was calculated to be 0.6% per year. The authors concluded that slippage was a rare finding overall in this population.^12^ In a small study of 40 Japanese patients aged 34 to 79 years with degenerative spondylolisthesis, progression greater than 5% slippage was observed in 30% with average radiographic observation over 9 years and 2 months. Mean % slippage or rate in change over time was not reported. The remaining 70% did not show progression with average observation over 7 years and 10 months.^13^ Slip progression seems to be most prevalent between the L4-L5 level according to a 15 year study; risk for slip progression was greatest for those between 40-60 years of age compared to those older than 60.^14^ In a group of 190 older men with average age of 74 years and average radiographic follow up of 4.6 years, prevalence of degenerative spondylolisthesis was 30% and out of this, only 12% had progression of spondylolisthesis. The data in this study gives an estimate of spondylolisthesis progression to be approximately 2.6% per year (95% CI 0.9-4.6%) in the 12% that had progression, though this study defined slippage as being greater than 5% and did not account for those that did not reach 5%.^11,15^

We present a study examining the effect of lumbar medial branch radiofrequency ablation (RFA) on patients with chronic lumbar pain originating from facets in the setting of Grade I or II lumbar spondylolisthesis. Facet-mediated pain may be present with degenerative spondylolisthesis through a presumed ventral-dorsal shearing stress on facets. RFA that denervates the innervation to lumbar facets may offer pain relief, but the spinal stabilizing effect of this therapy in such a population remains unknown. Medial branch nerves are targeted for thermal neurolysis in this therapy; however, ablation of adjacent branching nerves from the common dorsal ramus cannot always be avoided. These nerves are the intermediate and lateral branch nerves which provide innervation to the paraspinal muscles, longissimus and iliocostalis.^16^ The other major lumbar paraspinal muscles, multifidus, are innervated by the medial branch nerves after splitting off from branches innervating the facet joint.^17^ Transient muscle atrophy of the multifidus muscles after successful medial branch RFA has been demonstrated on MRI studies, though the long term clinical significance remains unclear. These muscles, with predominantly slow twitch fibers, serve as postural stabilizers of the spine.^18^ Standard thermal RFA lesioning at 80 degrees Celsius for 90 seconds with a standard 20- to 18-gauge monopolar radiofrequency needle has been shown to have a transverse distance of 5.3-5.9 millimeters, respectively. This may encompass branch nerves innervating paraspinal muscles, more so those innervating the multifidus muscle; however, no long term functional adverse effect has been demonstrated.^18,19^ It was our hypothesis that lumbar medial branch nerve RFA in patients with pre-existing degenerative spondylolisthesis would not result in advancement of listhesis post RFA observed over at least a 12-month period.

## Methods

In the course of usual treatment for lower back pain of facet etiology after exhausting conservative measures, 15 patients over age 40 with ongoing back pain and observed Grade I or Grade II spondylolisthesis who underwent or were planned to undergo lumbar medial branch nerve radiofrequency ablation were retrospectively or prospectively selected for observational study participation. Approval was obtained from the Institutional Review Board for Medical Ethics to recruit and treat patients who met the selection criteria. All participants had baseline imaging within 4 months of RFA in which there were clear lateral lumbosacral views for accurate measurement of baseline spondylolisthesis for comparison. Post-procedure imaging with lateral views through the lumbosacral spine was obtained at greater than 12 months. Of the 15 patients, one patient was not able to complete post-procedure imaging. A total of 14 patients completed the study.

Pre-procedure imaging included either those from the pre-RFA fluoroscopic diagnostic nerve block, fluoroscopic images from the RFA procedure, CT, MRI, or plain lumbar x-ray at baseline. Fluoroscopic images used were devoid of rotational artifact. Post-RFA lumbar x-rays with lateral views through the lumbosacral junction were obtained on all patients and funded from the study grant. However, if a CT or MRI was obtained for non-study purposes and exceeded 12 months post-ablation, this was used for spondylolisthesis measurement. This was done to minimize variability in spondylolisthesis due to weight-bearing changes between recumbent and standing position. All images were in neutral position and no dynamic imaging was performed. When fluoroscopic images were utilized for measurement, measurements were calibrated by comparing same-level vertebral body height to vertebral height on available radiographs, CTs, or MRIs. This was done to control for variable degrees of magnification inherent to the fluoroscopic modality. All participants were provided remuneration for the follow-up x-rays. Medial branch nerve radiofrequency ablation was performed by experienced interventional pain specialists using a parallel needle placement technique.^16^ Standard Stryker equipment including 18-gauge needles 100 to 150 mm in length with a standard 10-mm active tip was used and a standard thermal lesioning temperature of 80 degrees Celsius for 90 seconds was performed. All participants had symmetrical, bilateral RFA performed. All patients only had one RFA procedure performed during the study period but may have had previous ablation procedures prior to the study.

A single, experienced neuromuscular radiologist analyzed all pre- and post-RFA images and measured listhesis in millimeters based on the displacement of the caudad endplate of the superior vertebral level relative to the adjacent inferior vertebral level. A departmental biostatistician provided the statistical analysis.

### Statistical Methodology

A statistical power sensitivity analysis was conducted using G*Power 3.1.9.4.^20^ Performing a one-tailed, one-sample t-test with n=14, alpha=0.05 and power=0.80, a sample size of 14 is able to detect a “medium to large” effect size of 0.70. (“Medium” = 0.5, “Large” = 0.8).

Statistical analysis was performed by SAS 9.4 (Cary, NC). Data from 14 patients were obtained. The main outcome, percent advancement in listhesis per year, was calculated as: [(Post RFA – Pre RFA) / Endplate Distance] * 100% * (12 Months / Months Follow-up)

A positive number indicates listhesis advancement; a negative number indicates an improvement or measurement variability.

Comparisons of demographics and potential confounders against the outcome were performed with non-parametric tests, to avoid assumptions of normal distribution: Spearman Rank Correlation (instead of Pearson Correlation), Mann-Whitney U Test (instead of the t-test), and Kruskal-Wallis Test (instead of ANOVA). Because this study’s main hypothesis that there is no adverse effect of RFA on listhesis, we are effectively trying to prove the null hypothesis, and a traditional hypothesis test cannot be performed. Instead, we are performing a Method Comparison study, with the outcome descriptive statistics of the 95% confidence interval, compared against a suspected “natural” decline of about 2.6% as described by a study by Denard.^11^

### Results

Table 1 shows medians and interquartile range or frequencies for several demographic variables (age, gender, race, BMI). Using non-parametric statistical tests, there was no significant difference in percent advancement in listhesis per year by demographics: age, gender, race or BMI. There was also no significant difference by pre-RFA listhesis score, prior RFA, vertebrae involved or imaging technique. Additionally, using Spearman Rank Correlation, age was significantly negatively correlated with BMI (p=0.0029, r= −0.73) and follow-up time (p=0.029, r= −0.58). Post-RFA listhesis was positively correlated with pre-RFA listhesis (p=0.013, r=0.64) and percent advancement of listhesis per year (p=0.0086, r = 0.67).

**Table 1:** Demographics and Outcome Measure in % Advancement in Listhesis

| Continuous Variable | Median (IQR) |  | Mean (SD) | p-value |
| --- | --- | --- | --- | --- |
| BMI | 33.50 (29.5-36.4) |  |  | 0.53<sup>a</sup> |
| Age | 66 (55-70) |  |  | 0.099<sup>a</sup> |
| Pre-RFA Listhesis (mm) | 4.65 (3.7-5.8) |  |  | 0.90<sup>a</sup> |
| Post-RFA Listhesis (mm) | 5.00 (4.2-6.3) | | | $p=0.0086^a$<br>$r=0.67$ |
| Follow-up Time (months) | 23.50 (18-27) |  | 23.86 (9.03) | 0.15<sup>a</sup> |
| Percent advancement / year | 0.72 (-0.50 to 3.07) |  | 1.30 (2.55) | 0.12<sup>b</sup> |
| Categorical Variable | 1 | 2 | 3 | p-value |
| Gender | Female | Male |  | 0.60<sup>c</sup> |
|  | 9 (64.3%) | 5 (35.7%) |  |  |
| Race | African Am. | Caucasian | Unknown | 0.68<sup>c</sup> |
|  | 10 (76.9%) | 3 (23.1%) | 1 (NA) |  |
| Prior RFA | No | Yes |  | 0.90<sup>c</sup> |
|  | 9 (64.3%) | 5 (35.7%) |  |  |
| Vertebrae Listhesis | L4-L5 | L5-S1 |  | 0.44<sup>c</sup> |
|  | 9 (64.3%) | 5 (35.7%) |  |  |
| Imaging | L-spine MRI | L-spine X-Ray | Proc. Fluoro | 0.33<sup>d</sup> |
|  | 3 (21.4%) | 4 (28.6%) | 7 (50%) |  |
| Mismatched imaging pre- & post-RFA | No | Yes |  | 0.10<sup>c</sup> |
|  | 10 (71.4%) | 4 (28.6%) |  |  |

<sup>a</sup> Spearman Rank Correlation, equivalent to linear regression
<sup>b</sup> Signed Rank Test, equivalent to the one-sample t-test
<sup>c</sup> Mann-Whitney U Test, equivalent to the two-sample t-test
<sup>d</sup> Kruskal-Wallis Test, equivalent to ANOVA
Abbreviations: IQR: Interquartile Range; SD: Standard Deviation; RFA: Radio-frequency ablation

For the primary outcome, percent advancement of listhesis per year, 9 (64%) had values less than the expected 2.6 % without intervention, with the highest value of this subset 1.25%. Figure 1 shows the outcome by pre-RFA levels. Data for the outcome were slightly positively skewed, but multiple tests for normality showed it could be treated as normally distributed, including the Shapiro-Wilks Test (p=0.69). This yields a mean percent advancement of listhesis per year of 1.30% (95% CI −0.17 to 2.78), assuming normality.

**Figure 1:**
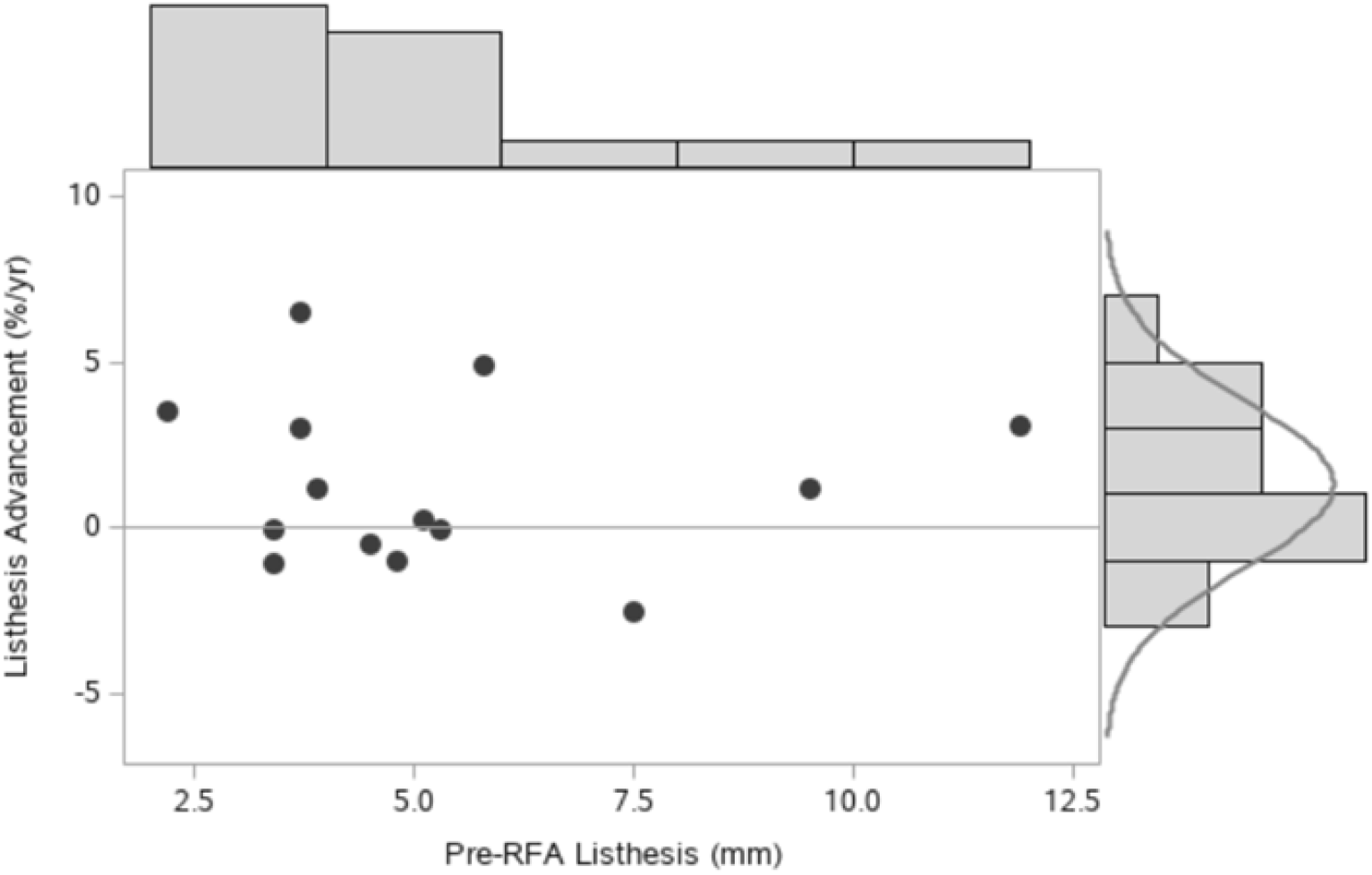
Scatterplot of the percent advancement in listhesis score per year as a function of the pre-RFA score. Positive numbers indicate an advancement of listhesis. Six patients (43%) saw a slight improvement in listhesis while an additional 3 patients (21%) showed advancement less than 1.25%, below the 2.6 %, the expected decline without intervention. Five patients (36%) saw an advancement of listhesis beyond 2.6%.

### Primary Outcome

The goal was to determine if there was advancement in listhesis measured in percent change per year over the observed 2.6 % per year baseline in the Denard study. Data for the percent change per year was normally distributed. No significant change was found (p=0.79). The mean change was 1.30% (95% CI −0.14 to 2.78%). Out of 14 patients, 8 (57%) had any advancement of listhesis over time (max: 70%), and 5 of the 8 had advancement greater than 2.6%. These patients had a median change of 3.48%, a minimum of 2.99% and a maximum of 6.50%. Out of 14 patients, 6 (43%) had slight reduction in listhesis (min: −12%). This study shows that performing RFA advances listhesis an average of less than 2.78% (upper limit of confidence interval).

## Discussion

The overall mean percent change of listhesis per year was 1.30% (95% confidence interval −0.17 to 2.78). Using the upper limit of the confidence interval, we observed a percent advancement of listhesis of up to 2.78% per year. This included patients who saw no change or a slight improvement in listhesis. One large observational study calculated the percent advancement of degenerative spondylolisthesis to be 2.6% per year (12%/4.6 years), but included those with greater than 5% slippage.^11^ This was observed in 12% of patients out of those with initial spondylolisthesis at the beginning of the observation period. We saw a progression in 5 patients out of 14 with baseline spondylolisthesis, with an assumption of a meaningful advancement occurring beyond 1.25% per year. Nine patients did not meet this level of progression, despite having had the RFA intervention. Looking at just the 5 patients with meaningful progression, we calculated a median progression of 3.48% (Range: 2.99-6.50%). We conclude that our findings are sufficiently similar to the natural progression of degenerative spondylolisthesis as best as we could define it, and the neuroablative procedure, lumbar medial branch nerve RFA does not destabilize the spine by advancing spondylolisthesis in this patient population. Some paraspinal muscle atrophy likely occurs post-procedure as previously shown, but is unlikely to contribute to further lumbar spine instability in the form of progressing degenerative spondylolisthesis.^18^

We did observe a slight decrease in spondylolisthesis over time in 6/14 (43%) of patients. We believe that this may have been mostly due to the small margin of error in measurement of listhesis that is on the order of millimeters. Several sources of error may occur to produce this: body position effects such as recumbency or rotational, variability in imaging resolution, or calibration across the different imaging modalities used. Improvement in spondylolisthesis may theoretically occur with strengthening of lumbar paraspinal and abdominal muscles as studies associate weakening or atrophy of these muscles to developing degenerative spondylolisthesis.^23,24^

Most patients in our study had greatest listhesis over L4-5 level which is consistent with previous epidemiological studies.^2,6,21^ Median BMI in our study was 33.5 which is categorized as obese. BMI as a risk factor for degenerative spondylolisthesis remains controversial.^6,7^ It has been theorized that excessive weight may exacerbate load and shearing forces on the spine and contribute to degenerative spondylolisthesis. Conversely, BMI may also contribute to degenerative disc disease, facet overlap with osteophytosis, and ossification of ligaments which may facilitate spondylolisthesis stabilization.^1,22^ The ratio of female: male in our study was 9:5. Though our sample size was too small to confirm the prevalence gender ratios reported in other studies, it is in accord with epidemiological studies reporting degenerative spondylolisthesis having a greater prevalence in women.^4,5,6^

### Study Limitations

This study has several limitations. First, the sample size is small. Hundreds of patients were screened through our center to identify patients with lumbar pain originating from facets undergoing RFA with coexisting spondylolisthesis and good quality baseline imaging within 4 months of RFA. Several international studies report a low prevalence of degenerative spondylolisthesis ranging from 12% up to 30%. This may be observed in the general American population as well and a small percentage will have pain, specifically of facet etiology. Second, our study is from a single institution. Current findings thus reflect on the clinical practices of the institution, which may limit the generalizability of the results. Third, in some patients, the pre- and post-RFA imaging was in different recumbency positioning which may affect observed spondylolisthesis to a small degree. This still remains controversial in the medical literature. Fourth, there remains limited data on the accepted rate of advancement of degenerative spondylolisthesis per year as a baseline comparison. We found a single study which analyzed such data, which can obfuscate comparison studies such as ours. Also, we are implicitly assuming a continuous linear progression of listhesis over time. In reality, the rate of degenerative spondylolisthesis progression may be quite variable with numerous contributing factors. Periods of peak progression have been described in prospective observational pediatric populations as well as older adult studies. Conversely, a slowing of progression or stabilization of spondylolisthesis has been described in the very elderly population. This was also observed in our study in which 9/14 with baseline spondylolisthesis did not show advancement of slippage beyond 1.25%. However, our median follow up period was 23.5 months, arguably not long enough to capture those with a very slow rate of progression of listhesis, assuming a somewhat continuous progression over time.

## Conclusion

Among patients with degenerative lumbar spondylosis with coexisting facetogenic pain who underwent medial branch nerve radiofrequency ablation, a mean spondylolisthesis advancement of 1.3% per year is comparable to the rate of natural progression without any intervention. Radiofrequency ablation in this patient population may be considered a safe therapy as a strategy for pain management. Larger studies with longer follow-up periods are needed to confirm this finding.

## Data Availability

All patient data is appropriated archived in a secured file and/or password encrypted manner.

